# Variation in SARS-CoV-2 seroprevalence in school-children across districts, schools and classes

**DOI:** 10.1101/2020.09.18.20191254

**Authors:** Agne Ulyte, Thomas Radtke, Irene A. Abela, Sarah R Haile, Jacob Blankenberger, Ruedi Jung, Céline Capelli, Christoph Berger, Anja Frei, Michael Huber, Merle Schanz, Magdalena Schwarzmueller, Alexandra Trkola, Jan Fehr, Milo A. Puhan, Susi Kriemler

## Abstract

**Importance:** Understanding transmission and impact of severe acute respiratory syndrome coronavirus 2 (SARS-CoV-2) in school children is critical to implement appropriate mitigation measures.

**Objective:** To determine the variation in SARS-CoV-2 seroprevalence in school children across districts, schools, grades, and classes, and the relationship of SARS-CoV-2 seroprevalence with self-reported symptoms.

**Design:** Cross-sectional analysis of baseline measurements of a longitudinal cohort study (Ciao Corona) from June-July 2020.

**Setting:** 55 randomly selected schools and classes stratified by district in the canton of Zurich, Switzerland (1.5 million inhabitants).

**Participants:** Children, aged 6-16 years old, attending grades 1-2, 4-5 and 7-8.

**Exposure:** Exposure to circulating SARS-CoV-2 between February and June 2020 including public lock-down and school closure (March 16-May 10, 2020).

**Main Outcomes and Measures:** Variation in seroprevalence of SARS-CoV-2 in children across 12 cantonal districts, schools, and grades using a Luminex-based antibody test with four targets for each of IgG, IgA and IgM. Clustering of cases within classes. Analysis of associations of seropositivity and symptoms. Comparison of seroprevalence with a randomly selected adult population, based on Luminex-based IgG and IgA antibody test of Corona Immunitas.

**Results:** In total, 55 schools and 2585 children were recruited (1337 girls, median age 11, age range 6-16 years). Overall seroprevalence was 2.8 % (95% CI 1.6–4.1%), ranging from 1.0% to 4.5% across districts. Seroprevalence was 3.8% (1.9-6.1%) in grades 1-2, 2.5% (1.1-4.2%) in grades 4-5, and 1.5% (0.5-3.0%) in grades 7–8. At least one case was present in 36/55 tested schools and in 43/128 classes with ≥50% participation rate and ≥5 children tested. 73% of children reported COVID-19 compatible symptoms since January 2020, but none were reported more frequently in seropositive compared to seronegative children. Seroprevalence of children was very similar to seroprevalence of randomly selected adults in the same region in June-July 2020, measured with the same Corona Immunitas test, combining IgG and IgA (3.1%, 95% CI 1.4-5.4%, versus 3.3%, 95% CI 1.4-5.5%).

**Conclusions and Relevance:** Seroprevalence was inversely related to age and revealed a dark figure of around 90 when compared to 0.03% confirmed PCR+ cases in children in the same area by end of June. We did not find clustering of SARS-CoV-2 seropositive cases in schools so far, but the follow-up of this school-based study will shed more light on transmission within and outside schools.

**Trial registration:** http://ClinicalTrials.gov Identifier: NCT04448717, registered June 26, 2020. https://clinicaltrials.gov/ct2/show/NCT04448717

**Key Points:** *Question:* What is the variation in seroprevalence of SARS-CoV-2 cases in school children across districts, schools and classes?

*Findings:* Among 55 randomly selected schools and 2585 children, 2.8 % (95% CI 1.6-4.1%) of children had SARS-CoV-2 antibodies, similar to the seroprevalence in the adult population in the same region but showing a much higher dark figure (89 versus 12 in adults). A third of tested classes had at least one SARS-CoV-2 case, and higher seroprevalence was observed in lower grades. Seropositive children did not report SARS-CoV-2 infection symptoms more often than seronegative children.

*Meaning:* The results so far do not suggest substantial transmission within schools. In contrast to the current literature, younger children seem to be infected slightly more often; the striking dark figure of 89 could be partly due to the fact that symptoms compatible with COVID - 19 were highly prevalent in children and do not help to differentiate between seropositive and seronegative children.

## Introduction

The transmission of SARS-CoV-2 in school setting is not well understood^1^ – partly as schools were closed in many countries during the peaks of the pandemic, partly due to lack of representative studies with random sampling. Anecdotal evidence suggests that outbreaks can happen in schools^2^, and camping facilities^3^, but it is not clear if they represent outlier events or widely underdiagnosed spread of the infection.

In this study, we present the results of the first cross-section of a cohort of children from randomly selected schools and classes in the canton of Zurich, Switzerland, enrolled from June 16 to July 9, 2020. Schools in Switzerland were closed for a relative short period (March 16 to May 10) compared to other countries, and lock-down measures were mild. The cohort study follows the seroprevalence, symptoms, sociodemographic and life-style factors of enrolled children from June 2020 to April 2021.

The aim of this analysis is to present the overall estimate of seroprevalence and its variation within districts, schools, grades and classes, and the association of seroprevalence with symptoms.

## Methods

The protocol for this longitudinal cohort study is reported elsewhere^4^. The study is part of a large nationally coordinated research network Corona Immunitas in Switzerland^5,6^. A random sample of schools, stratified by district, and randomly selected classes in lower school level (grades 1-2, attended by 6 to 9-year-old children), middle (grades 4-5, 9 to 13-year-old children), and upper school level (grades 7-8, 12 to 16-year-old children), ensuring that the same cohort of children within the class can be followed until April 2021. We aimed to enroll at least 3 classes and 60 children per school level. Children were enrolled and venous blood samples taken in schools between June 16 and July 9. Questionnaires with information on socio-demographics and SARS-CoV-2 infection symptoms were completed online by parents. Information on schools was collected from school principals and the Educational Department of the canton of Zurich. The canton of Zurich comprises 1.5 million residents, roughly 18% of the Swiss population, and includes both urban and rural settings, as well as an ethnically and linguistically diverse population.

In total, 55 out of 156 invited primary and secondary schools agreed to participate, and 2585 children were enrolled. Venous blood samples were collected from 2484 children, and online questionnaire completed for 2259 children. Blood samples were analyzed with a binding assay of the Institute of Medical Virology (IMV) of the University of Zurich based on the Luminex technology. The test analyzes immunoglobulins G, M and A against four SARSCoV-2 targets (receptor binding domain (RBD), spike proteins S1 and S2, and the nucleocapsid protein (N), yielding 12 different measurements. Cut-off values were established against pre-pandemic plasma allowing a high sensitivity (93.3%) and specificity (99.6%). Samples were defined as seropositive for SARS-CoV-2 if at least two of the 12 parameters were above cut-off.

SARS-CoV-2 seroprevalence in children was compared to that estimated in a random sample from the general population, adjusting for age group and sex, in the same region in June-July 2020. The adult study, as all studies of the Swiss-wide research program Corona Immunitas^5,6^, used the test SenASTrIS (Sensitive Anti-SARS-CoV-2 Spike Trimer Immunoglobulin Serological) developed by the Centre Hospitalier Universitaire Vaudois (CHUV), the Swiss Federal Institute of Technology in Lausanne (EPFL) and the Swiss Vaccine Center^7^. The test also uses Luminex technology to detect immunoglobulin G (IgG) and IgA antibodies binding to the entire trimeric S protein of SARS-CoV-2 and with demonstrated 98.3% sensitivity and 98.4% specificity for IgG and IgA combined. In order to compare the seroprevalence estimates in children and adult cohorts, a sample of 1717 collected children blood samples was also analyzed with the SenASTrIS test and compared to a random sample of 577 adults who take part in the Corona Immunitas research program^6^.

Seroprevalence was also compared to the cumulative incidence of reverse transcription polymerase chain reaction (RT-PCR)-confirmed SARS-CoV-2 infections in adults and children, based on official statistics up to beginning of June^8^.

Statistical analysis included descriptive statistics and Bayesian hierarchical modelling to estimate seroprevalence, accounting for the sensitivity and specificity of the SARS-CoV-2 antibody test, the hierarchical structure of cohort (individual and school levels), and post-stratification weights, which adjusted for population-level grade level and geographic district^9^.

## Results

In total, 55 schools and 2585 children were recruited (1337 girls, median age 11, age range 6–16 years), 754 in lower level, 899 in middle, and 932 in upper school level. Mean participation rate across classes was 45%, ranging from 5% to 94% (1 to 21 participating children). Venous blood was collected and analyzed for 2484 children (1276 girls, median age 11, age range 6-6 years).

74 children had SARS-CoV-2 antibodies, resulting in overall weighted seroprevalence of 2.8 % (95% credible interval 1.6-4.1%), ranging from 1.0% to 4.5% in districts (Figure 1). Seroprevalence was 3.8% (1.9-6.1%) in grades 1-2, 2.5% (1.1-4.2%) in grades 4-5, and 1.5% (0.5-3.0%) in grades 7-8 (Figure 1).

**Figure 1.**
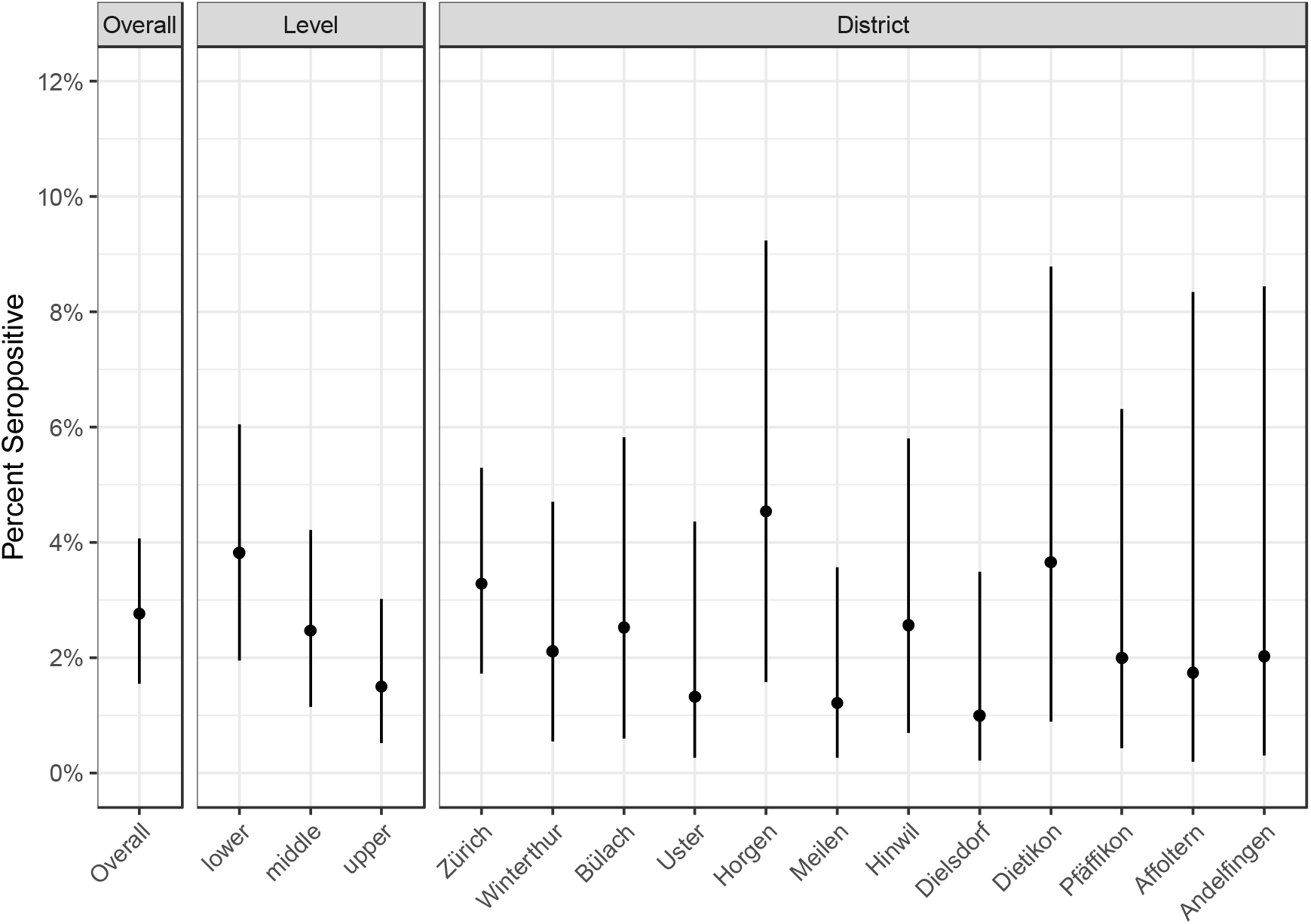
Overall, school level and district estimates of SARS-CoV-2 seroprevalence in children

Weighted point estimates and 95% credible intervals are shown. Districts are ordered by population size.

Seroprevalence of children, as measured with the Corona Immunitas test of IgG and IgA combined, was very similar to seroprevalence of randomly selected adults in the same region in June-July 2020 (3.1%, 95% CI 1.4-5.4%, versus 3.3%, 95% CI 1.4-5.5%). The other estimates based on the Corona Immunitas test in the children cohort were similar to those reported above, in different age groups, sexes, and districts.

Based on SARS-CoV-2 RT-PCR-confirmed cases by the end of June (0.03% for children and 0.24% for adults), the factor of confirmed to total infections (dark figure) in children was 89, compared to a factor of 12 in the adult Swiss population.

At least one seropositive child was present in 36/55 of tested schools. 34% (43/128) of classes, with ≥5 children and ≥50% of children within class tested, had at least one seropositive case (Figure 2). When considering all classes regardless of participation rate, 24% of classes had at least one case; whereas when considering higher inclusion threshold of ≥15 children and ≥60% of the class tested, 45% of classes had at least one case.

**Figure 2.**
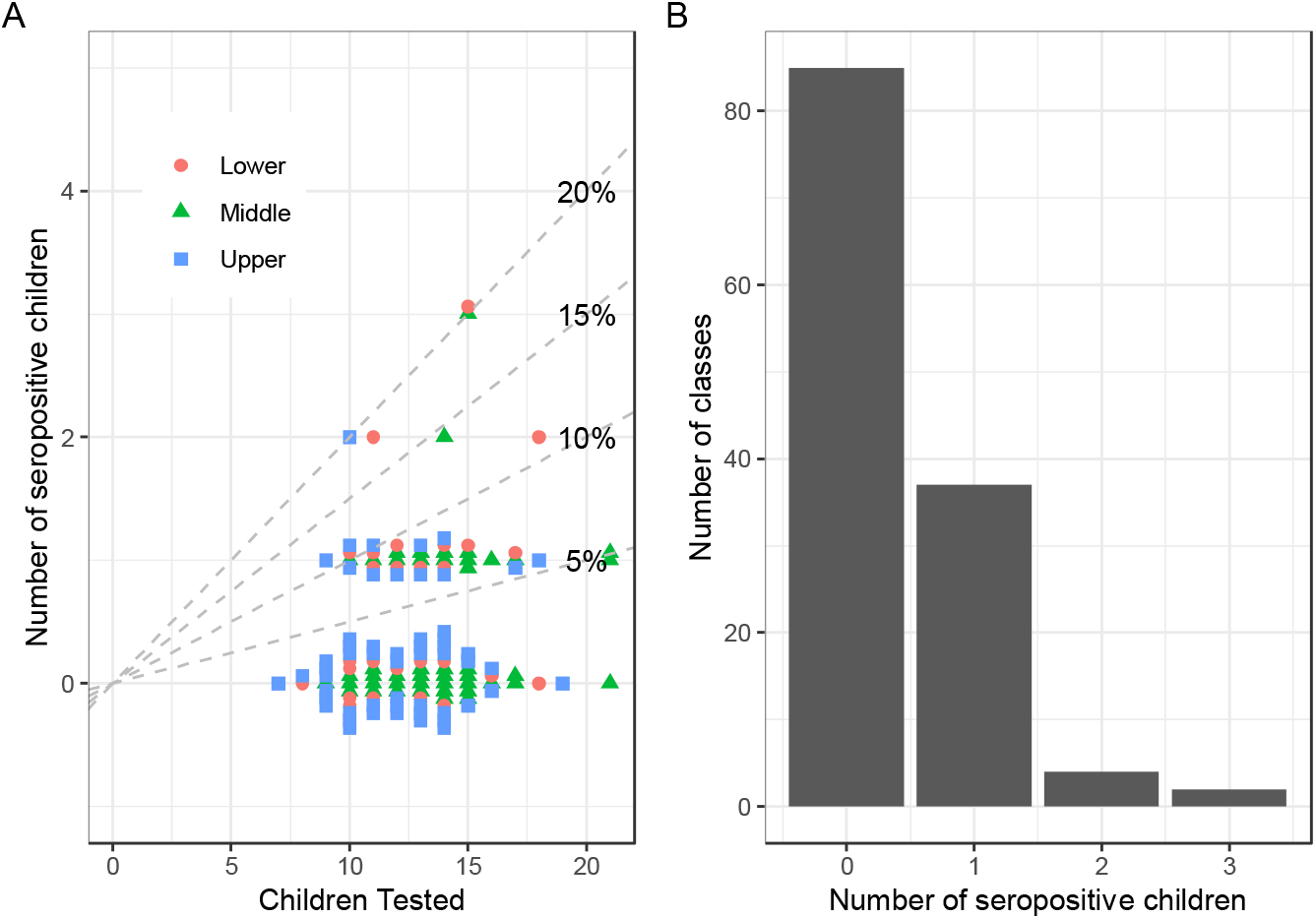
Clustering of seropositive children in classes

Only classes where at least 5 children and at least 50% of the class were tested are shown.

No sex differences in seroprevalence were noted (2.8% (1.6-4.1%) in girls and 2.7% (1.5-4.0%) in boys). 73% of children reported any SARS-CoV-2 compatible symptoms between January and June 2020. None of the symptoms were more frequent in seropositive than in seronegative children (Figure 3).

**Figure 3.**
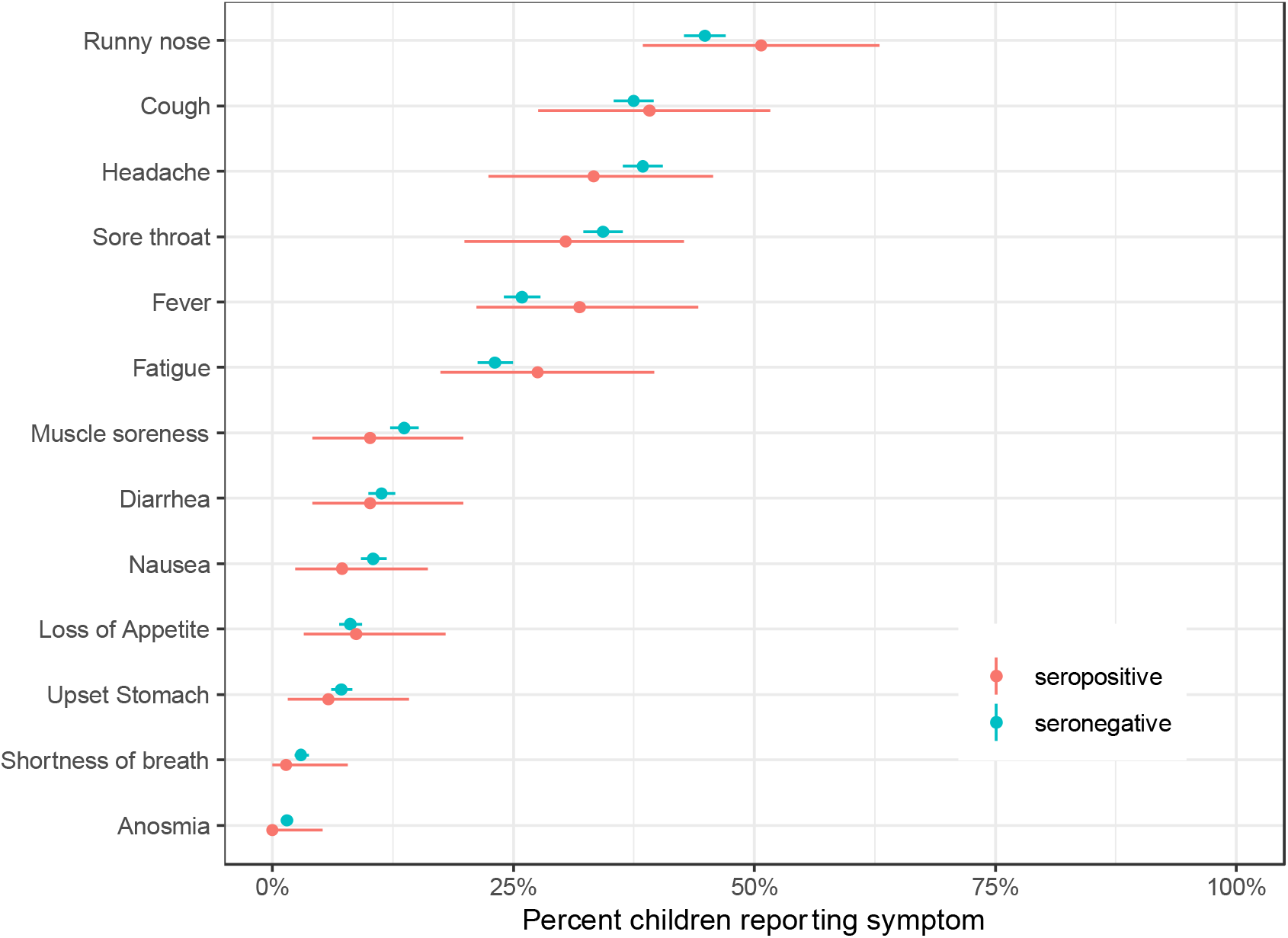
Self-reported symptoms in seropositive and seronegative children in January-June 2020

Point estimates and 95% credible intervals are shown.

## Discussion and conclusions

In this study of randomly sampled schools and classes by July 2020, thus covering the period between February to June 2020, we found variation in seroprevalence in 6 to 16-year-old children across districts, schools and classes, but no indication of major transmission within schools. The overall seroprevalence was not different from that in adults – pointing to striking underdiagnosis of SARS-CoV-2 infection in children, with only 1 in 90 cases diagnosed. Such a high dark figure is likely explained by our observation that no symptoms seem to be suggestive of a SARS-CoV-2 infection in children, contrary to more specific symptoms in adults^10^, and by cautious testing indications by health authorities. Contrary to studies of symptomatic infections^11^, there was a trend of higher seroprevalence in younger children. Although no outbreaks were reported in schools at the time of testing in the canton of Zurich (comprising 18% of the Swiss population), seropositive cases were detected in more than half of the tested schools and a third of all tested classes. The vast majority of classes with cases had only a single case. The presence of symptoms was very common (3 of 4 children reported one or several symptoms compatible with a COVID-19 infection) and importantly not specific to the seropositive children.

The management of SARS-CoV-2 transmission in schools is highly debated^12,13^, as current evidence consists mostly of case studies and series of individual schools, reporting mostly low secondary attack rates in schools^14,15^, but also conflicting observations of outbreaks^2,3^. Although some studies of seroprevalence have included children^9,16^, they mostly focused on households and the general population. The present study is thus unique as one of the first major studies reporting variation in seroprevalence in children from randomly selected schools in a country where the general lock-down on a population level was mild and short (one month), and school closure lasted only for two months.

Although manifest clinical disease of COVID-19 is much less prevalent in children than adults^11^, we observed a trend of younger children having higher seroprevalence than older children. This could be related to virtually impossible social distancing behavior in younger children or a different immune response to the virus. The symptomatology was not different among seropositive and seronegative children, with 3 of 4 children reporting symptoms within the last 6 months in both groups. Specificity of COVID-19 symptoms therefore seems to be lower in children than in adults.

In conclusion, clustering of SARS-CoV-2 seropositive cases within schools and grades was not prominent shortly after reopening of schools in this population-based study. Seroprevalence was similar to adults and higher in lower grade compared to higher grade children, resulting in a strikingly higher dark number than in adults, in particular in the very young children. Considering the time window required for SARS-CoV-2 antibodies to form, this study reflects infection of SARS-CoV-2 until approximately end of May 2020, covering a period of four months of SARS-CoV-2 infection in the community, with two months of school closure and mild lock-down policy. The subsequent testing of parents and school personnel and the follow-up of the children cohort in fall 2020 will yield further evidence of the spread of SARS-CoV-2 within and outside schools.

## Data Availability

Data is currently not available due to participant privacy constraints. Eventual availability of data is under consideration.

## Acknowledgements

Acknowledgements

## Authors’ Contributions

SK and MAP initiated the project and preliminary design, with support of JF. SK, MAP, CB, TR, RJ, JB, AF and AU developed the design and methodology. SK, RJ, AU, TR, JB, AF and CC recruited study participants, collected and managed the data. SH performed statistical analysis. AT, MH, MaSch, MeSch and IA developed the serology analysis plan, supervised, conducted and evaluated the serology tests. AU wrote the first draft of the manuscript. All authors contributed to the design of the study and interpretation of its results, and revised and approved the manuscript for intellectual content.

## Author access to data

TR had full access to all the data in the study and takes responsibility for the integrity of the data and the accuracy of the data analysis. SK, TR, AU and MAP had access to full study data. AU, TR, JB and CC were primarily responsible for data management. SH was primarily responsible for data analysis.

## References

1. Viner RM, Russell SJ, Croker H, et al. School closure and management practices during coronavirus outbreaks including COVID-19: a rapid systematic review. Lancet Child Adolesc Heal. 2020; 4(5): 397-404. doi: 10.1016/S2352-4642(20)30095-X

2. Stein-Zamir C, Abramson N, Shoob H, et al. A large COVID-19 outbreak in a high school 10 days after schools’ reopening, Israel, May 2020. Eurosurveillance. 2020; 25(29): 2001352. doi: 10.2807/1560-7917.ES.2020.25.29.2001352

3. Szablewski CM, Chang KT, Brown MM, et al. SARS-CoV-2 Transmission and Infection Among Attendees of an Overnight Camp — Georgia, June 2020. MMWR Morb Mortal Wkly Rep. 2020; 69(31): 1023–1025. doi: 10.15585/mmwr.mm6931e1

4. Ulyte A, Radtke T, Abela I, et al. Seroprevalence and immunity of SARS-CoV-2 infection in children and adolescents in schools in Switzerland: design for a longitudinal, school-based prospective cohort study. medRxiv. September 2020: 2020. 08.30.20184671. doi: 10.1101/2020.08.30.20184671

5. Corona Immunitas: a nationwide program of antibody studies of SARS-CoV-2 in the Swiss population (ISRCTN18181860). http://www.isrctn.com/ISRCTN18181860. Accessed August 28, 2020.

6. West EA, Anker D, Amati R, et al. Corona Immunitas: study protocol of a nationwide program of SARS-CoV-2 seroprevalence and seroepidemiologic studies in Switzerland. Int J Public Health. 2020; In press.

7. Fenwick C, Croxatto A, Coste AT, et al. Changes in SARS-CoV-2 Antibody Responses Impact the Estimates of Infections in Population-Based Seroprevalence Studies. medRxiv. July 2020: 2020.07.14.20153536. doi: 10.1101/2020.07.14.20153536

8. Canton of Zurich. Numbers and Facts on COVID-19 [Kanton Zürich. Zahlen & Fakten zu COVID-19]. https://www.zh.ch/de/gesundheit/coronavirus/zahlen-fakten-covid-19.html?keyword=covid19#/home. Accessed August 28, 2020.

9. Stringhini S, Wisniak A, Piumatti G, et al. Seroprevalence of anti-SARS-CoV-2 IgG antibodies in Geneva, Switzerland (SEROCoV-POP): a population-based study. Lancet. 2020; 0(0). doi: 10.1016/S0140-6736(20)31304-0

10. Struyf T, Deeks JJ, Dinnes J, et al. Signs and symptoms to determine if a patient presenting in primary care or hospital outpatient settings has COVID-19 disease. Cochrane Database Syst Rev. 2020;2020(7). doi: 10.1002/14651858.CD013665

11. de Lusignan S, Dorward J, Correa A, et al. Risk factors for SARS-CoV-2 among patients in the Oxford Royal College of General Practitioners Research and Surveillance Centre primary care network: a cross-sectional study. Lancet Infect Dis. 2020; 20(9): 1034–1042. doi: 10.1016/S1473-3099(20)30371-6

12. Esposito S, Principi N. School Closure during the Coronavirus Disease 2019 (COVID-19) Pandemic: An Effective Intervention at the Global Level? JAMA Pediatr. 2020. doi: 10.1001/jamapediatrics.2020.1892

13. Christakis DA. School Reopening-The Pandemic Issue That Is Not Getting Its Due. JAMA Pediatr. 2020. doi: 10.1001/jamapediatrics.2020.2068

14. Heavey L, Casey G, Kelly C, Kelly D, McDarby G. No evidence of secondary transmission of COVID-19 from children attending school in Ireland, 2020. Eurosurveillance. 2020; 25(21): 2000903. doi: 10.2807/1560-7917.ES.2020.25.21.2000903

15. Macartney K, Quinn HE, Pillsbury AJ, et al. Transmission of SARS-CoV-2 in Australian educational settings: a prospective cohort study. Lancet Child Adolesc Heal. 2020; 0(0). doi: 10.1016/s2352-4642(20)30251-0

16. Pollán M, Pérez-Gómez B, Pastor-Barriuso R, et al. Prevalence of SARS-CoV-2 in Spain (ENE-COVID): a nationwide, population-based seroepidemiological study. Lancet (London, England). 2020; 0(0). doi: 10.1016/S0140-6736(20)31483-5

